# Evaluation of the Cepheid Xpert Xpress SARS-CoV-2 test for bronchoalveolar lavage

**DOI:** 10.1101/2021.12.21.21268190

**Authors:** Tung Phan, Ashley Mays, Melissa McCullough, Alan Wells

**Author notes:** Correspondence to: Tung Phan MD, PhD. Department of Pathology, University of Pittsburgh, Pittsburgh, PA 15213, USA.

## Abstract

Accurate and rapid laboratory tests are essential for the prompt diagnosis of COVID-19, which is important to patients and infection control. The Xpert Xpress SARS-CoV-2 test is a real-time RT-PCR intended for the qualitative detection of nucleic acid from SARS-CoV-2 in upper respiratory specimens. In this study, we assessed the analytical and clinical performance characteristics of this rapid test for SARS-CoV-2 in 60 bronchoalveolar lavage (BAL) specimens. BAL is a specimen type that is not authorized under EUA for the Xpert Xpress SARS-CoV-2 test. The limit of detection of the Xpert Xpress SARS-CoV-2 test was 500 copies/ml. The overall agreement of the Xpert Xpress SARS-CoV-2 test was 100%. The Xpert Xpress SARS-CoV-2 test is sensitive and specific to aid in diagnosis of COVID-19 using bronchoalveolar lavage.

## The study

Coronaviruses are important human and animal pathogens associated with a variety of diseases from mild conditions to very severe illnesses that require hospitalization, intensive care unit admission and mechanical ventilation [1, 2]. There have been tireless efforts to understand virology, transmission and pathogenesis of SARS-CoV-2 and the management of COVID-19; however, the ongoing COVID-19 pandemic continues to pose significant challenges to public health beyond the impact of the disease itself [3, 4]. The United States is one of the emerging epicenters of the COVID-19 pandemic and surpassed 800,000 deaths from SARS-CoV-2 (https://coronavirus.jhu.edu/map.html). Diagnostic testing availability and capacity are the critical components to meet the challenge of the COVID-19 pandemic. Among many commercial diagnostic tests approved under the FDA’s emergency use authorization (EUA) process, nucleic acid amplification tests (NAATs) are the primary mean in diagnosing COVID-19 and they are the gold standard diagnostic method [5, 6]. NAATs are developed based on genomic information of SARS-CoV-2. Different tests have different genetic targets including the envelope gene (E), the nucleocapsid gene (N), the ORF1ab gene (RdRp) and the spike gene (S). Most NAATs utilize a dual- or triple-target design to safeguard against potential mutations in the SARS-CoV-2 genome [7, 8].

The Xpert Xpress SARS-CoV-2 test is a real-time RT-PCR authorized by the FDA under emergency use authorization (EUA) for the qualitative detection of nucleic acid from SARS-CoV-2 in upper respiratory specimens such as nasopharyngeal swab, nasal swab or nasal wash (https://www.cepheid.com/coronavirus). However, lower respiratory specimens (including bronchoalveolar lavage) are not authorized under the EUA for the Xpert Xpress SARS-CoV-2 test. This molecular test is designed to detect both E and N2 genes, and it can provide rapid detection of SARS-CoV-2 as soon as thirty minutes for positive results with less than a minute of hands-on-time to prepare clinical specimens (https://www.cepheid.com/coronavirus). The Xpert Xpress SARS-CoV-2 test is a fully integrated sample-to-answer assay, and it has been demonstrated to be highly sensitive and specific [9, 10]. Given the major advantages of this test along with bronchoalveolar lavage (BAL) being widely accepted in clinical settings, the evaluation of the Xpert Xpress SARS-CoV-2 test to detect SARS-CoV-2 in BALs is essential.

In this evaluation study, we used the GeneXpert Infinity System (Cepheid, Sunnyvale, CA), which is an automated molecular device performing specimen processing and real-time RT-PCR. The analytical sensitivity of the Xpert Xpress SARS-CoV-2 test was performed using one lot of testing cartridges, reagents, and serial dilutions of the quantitated SARS-CoV-2 (ZeptoMetrix). The limit of detection (LOD) was assessed by analyzing SARS-CoV-2 simulated BAL specimens with known viral load ranging from 125 to 1,000 copies/ml. All specimen dilutions were prepared using a clinical negative BAL matrix. For each dilution, 300 µL was tested using the Xpert Xpress SARS-CoV-2 test as per the manufacturer’s instructions. The LOD titer is defined as the lowest concentration at which ≥95% of specimens tested generated positive calls. A minimum of 20 replicates were tested for the LOD verification. Table 1 showed that the positivity rate of 20 replicates observed was ≥95% at 500 copies/ml for both N2 (20/20, 100%) and E (19/20, 95%) targets. Since FDA requires claimed LOD based on the least sensitive target, the LOD of the Xpert Xpress SARS-CoV-2 test for BAL was determined as 500 copies/ml.

**Table 1:** Limit of detection of the Xpert Xpress SARS-CoV-2 test to detect SARS-CoV-2 in bronchoalveolar lavage

| Copies/ml | E gene (% positive) | N2 gene (% positive) |
| --- | --- | --- |
| 1,000 | 5/5 (100%) | 5/5 (100%) |
| 500 | 19/20 (95%) | 20/20 (100%) |
| 250 | 4/10 (40%) | 10/10 (100%) |
| 125 | 1/5 (20%) | 5/5 (100%) |

To assess clinical performance characteristics of the Xpert Xpress SARS-CoV-2 test for BAL, a total of 60 BAL specimens (30 residual clinical negatives, 6 residual clinical positives, and 24 SARS-CoV-2 simulated positives) were tested using the Xpert Xpress SARS-CoV-2 test. The SARS-CoV-2 simulated specimens were made by spiking SARS-CoV-2 (ZeptoMetrix) in a clinical negative BAL matrix to generate 6 different levels from 1X LOD to 30X LOD. Results from the Xpert Xpress SARS-CoV-2 test were compared to the original results from the CDC 2019-nCoV RT-PCR Diagnostic Panel, which served as the reference method (https://www.cdc.gov/coronavirus/2019-ncov/lab/testing.html). In a side-by-side comparison between 30 positive BAL specimens and 30 negative BAL specimens, there was 100% agreement between the Xpert Xpress SARS-CoV-2 test and the CDC 2019-nCoV RT-PCR Diagnostic Panel. The clinical sensitivity was 100% (30/30), and the clinical specificity was 100% (30/30) as shown in Table 2.

**Table 2:** Clinical sensitivity, specificity, and agreement of the Xpert Xpress SARS-CoV-2 test to detect SARS-CoV-2 in bronchoalveolar lavage

| Xpert Xpress SARS-CoV-2 test | Reference |  |
| --- | --- | --- |
|  | Positive | Negative |
| Positive | 30 | 0 |
| Negative | 0 | 30 |
| Sensitivity | 30/30=100% |  |
| Specificity | 30/30=100% |  |
| Agreement | (30+30)/(30+30)=100% |  |

While routine bronchoscopy with BAL is not recommended as the first procedure to detect SARS-CoV-2 due to a risk of aerosolization, the detection of SARS-CoV-2 in BAL can significantly impact patient care in appropriate clinical scenarios. Gualano and colleagues reported an interesting case of a patient with a very high suspicion for COVID-19-associated pneumonia. The patient had two negative SARS-CoV-2 upper respiratory tract specimens, and then the virus was detected in BAL [11]. A recent study reported that SARS-CoV-2 was detected in 16% of 198 BAL specimens collected from the COVID-19 suspected patients who had single, double, or triple negative nasopharyngeal swab before bronchoscopy was performed [12]. In this study, our evaluation revealed that the Xpert Xpress SARS-CoV-2 test performed similarly to the CDC 2019-nCoV RT-PCR Diagnostic Panel on the BAL specimens. There were not any discrepant results between these two molecular assays. We did not see any false positives or false negatives when tested the negative or positive BAL specimens, respectively. There were not any failures of sample processing control (SPC), which is present to control for adequate processing of the specimens and to monitor for the presence of potential inhibitors in the RT-PCR reaction. In addition, invalid or canceled results were not observed during the evaluation. Precision analysis was also performed by testing two negative and two positive BAL specimens, which were run by 3 different technologist, and all of them (100%) yielded expected results. According to the Cepheid’s package insert, if both targets (E and N2) are detected or if only N2 is detected, the test reports a positive result. If only E is detected, the test reports a presumptive positive result because the target is shared with other coronaviruses belonging to the *Sarbecovirus* subgenus of the *Coronaviridae* family. While the LOD of the Xpert Xpress SARS-CoV-2 test for BAL is 500 copies/ml, the test can detect N2 at the lower concentration of 125 copies/ml as shown in Table 2, indicating that the test can report a positive result even there is a low viral quantity in the specimen. Taken together, the Xpert Xpress SARS-CoV-2 test effectively detects SARS-CoV-2 RNA directly from BAL, demonstrating high sensitivity, specificity, and accuracy.

## Data Availability

All data produced in the present work are contained in the manuscript

## Acknowledgements

We thank the UPMC Clinical Microbiology Laboratory for testing the specimens and performing the evaluation.

## Author contributions

TP and AW: designed the study and wrote the manuscript; AM and MM: managed the testing.

## Ethical approval

All testing was performed as apart of routine clinical care and performed according to CLIA ‘88 regulations by appropriate personnel. The entire study was deemed to be a Quality Improvement initiative by the UPMC IRB and approved by the UPMC QI Review Board.

